# Differential expression of plasma proteins in pregnant women exposed to *Plasmodium falciparum* malaria

**DOI:** 10.1101/2024.05.01.24306614

**Authors:** Bernard N. Kanoi, Harrison Waweru, Francis M. Kobia, Joseph Mukala, Peter Kirira, Dominic Mogere, Radiosa Gallini, Mikael Åberg, Manu Vatish, Jesse Gitaka, Masood Kamali-Moghaddam

**Author notes:** **Correspondence:** Centre for Malaria Elimination, Institute of Tropical Medicine, Mount Kenya University, Thika, Kenya. Bernard Kanoi, Centre for Malaria Elimination, Institute of Tropical Medicine, Mount Kenya University, Thika, Kenya. Jesse Gitaka, ^4^Department of Immunology, Genetics & Pathology, Science for Life Laboratory, Uppsala University, Uppsala, Sweden, Masood Kamali-Moghaddam. Shared senior authorship (Jesse Gitaka, Masood Kamali-Moghaddam).

## Abstract

In sub-Saharan Africa, pregnant women are at greater risk of malaria infection than non-pregnant adult women. The infection may lead to pregnancy-associated malaria (PAM) because of the sequestration of *Plasmodium falciparum*-infected erythrocytes in the placental intervillous space. Although there are several tools for diagnosing malaria infection during pregnancy, including blood smear microscopic examination, rapid diagnostic tests, and PCR, there are no tools for detecting placental infection and, by extension, any dysfunction associated with PAM. Thus, PAM, specifically placental infection, can only be confirmed via postnatal placental histopathology. Therefore, there is an urgent need for specific serum biomarkers of PAM. Here, we used the high throughput proximity extension assay to screen plasma from malaria-exposed pregnant women for differentially expressed proteins that can predict PAM or adverse malaria outcomes. Such biomarkers may also elucidate the pathophysiology of PAM. We observed that the IgG Fc receptor IIb (Uniprot ID P31994) and HO-1 (P09601) are consistently highly expressed in malaria-positive samples compared to samples from malaria-negative pregnant women. On the contrary, NRTN (Q99748) and IL-20 (Q9NYY1) were differentially expressed in the malaria-negative women. IL-20 exhibited the highest discriminatory power (AUC = 0.815), indicating a strong association with malaria status. These proteins should be considered for further evaluation as biomarkers of malaria-induced placental dysfunction in pregnant women.

## Introduction

Pregnant women in most tropical areas are at an increased risk of malaria because of the placental sequestration of *Plasmodium falciparum* (*P. falciparum*)-infected red blood cells, which can lead to stillbirth and low birth weight [1]. Despite the availability of numerous diagnostic tools for detecting malaria during pregnancy, there is a lack of a definitive test for determining actual placental infection and dysfunction within the context of pregnancy-associated malaria (PAM) [2]. PAM has several similarities with pre-eclampsia, for which several biomarkers of placental stress and dysfunction have been identified [3,4]. While some of these biomarkers are inflammatory markers that lack specificity, others, such as sendoglin and sFlt1, which are elevated in peripheral blood during pre-eclampsia, appear more specific to placental dysfunction [5]. The predictive value of such biomarkers should be evaluated in the context of PAM to determine their effectiveness as indicators of PAM-associated placental infection and dysfunction [6]. This is crucial for enhancing the diagnosis of typically sub-microscopic malaria infections and malaria-induced placental dysfunction, thereby facilitating timely intervention in pregnant women. Indeed, comprehension of disease biology and the interaction between the malaria parasite and the human host is crucial for developing diagnostic, prognostic, and uncovering predictive biomarkers for PAM [7]. Elucidating the expression patterns of plasma proteins can provide insights into how PAM affects placental, maternal, or fetal wellbeing [6,8].

The rapid development of proteomics platforms, such as multiplex proximity extension assay (PEA), offers highly effective, high-throughput strategies for studying malaria-associated molecular pathophysiological changes [9]. PEA technology enables simultaneous measurement of a large number of proteins using minute amount of sample [7,10]. Studies using this approach have demonstrated its clinical utility in identifying important biomarkers for various diseases, including cancer, long COVID, ischemic stroke, and early pregnancy bleeding [10–15]. In this study, we used PEA to screen sera from pregnant, malaria-exposed women to identify differentially expressed proteins. Using this strategy, we identified four proteins significantly differentially expressed in samples from malaria-infected pregnant women compared with non-infected pregnant counterparts, suggesting that their alteration is *P. falciparum-*driven. Such proteins can potentially develop into biomarkers of PAM or adverse malaria outcomes.

## Material and methods

### Study design & population

This study used serum samples collected from women between 12 and 18 weeks of gestation. These women were enrolled in the study during their routine antenatal clinic visits at Webuye County Hospital in Bungoma County, Kenya, a region characterized by high malaria transmission [16]. The study protocol and the use of this well-characterized biobank of samples were approved by the Independent Ethics Research Committee (IERC) of Mount Kenya University (No# MKU/IERC/0543, MKU/IERC/2461). The enrolled cohort consisted of women undergoing routine intermittent preventive malaria treatment in pregnancy (IPTp) with sulfadoxine-pyrimethamine (IPTp-SP) [17]. All study participants gave written informed consent before joining the study. The study protocol complied with the International Conference on Harmonization Good Clinical Practices and the Declaration of Helsinki guidelines [18]. Serum samples were collected 4 weeks after IPTp administration and frozen immediately, and then transported on dry ice to the Centre for Malaria Elimination laboratories at Mount Kenya University and immediately stored in a -80°C deep freezer without cold-chain interruption. Each serum sample was matched with corresponding anonymized demographic and pregnancy outcome data. Data on pregnancy progress and outcomes were collected during the scheduled and unscheduled visits. Malaria diagnosis was performed using both microscopy and a ParaCheck Rapid Diagnostic Kit (Orchid Biomedical Systems, India).

### Protein abundance measurement by multiplex proximity extension assay

Multiplex PEA technology was used to analyze protein expression in the sera samples using the Olink Inflammation and Cardiovascular II protein panels (Olink™ Proteomics, Uppsala Sweden, https://olink.com), loading 1µL sample/panel. These panels were selected since they target inflammatory and vascular pathophysiological proteomic signatures, which were hypothesized to be altered by PAM. In PEA, each target protein is recognized by two antibodies attached to single-stranded DNA oligonucleotide [11]. When a pair of antibodies binds to the same target protein, the conjugated DNA oligonucleotides are brought in proximity and hybridize to each other. Subsequently, the DNA molecules are extended to form a double-stranded DNA as template for quantitative PCR amplification [11]. The results are presented as normalized protein expression (NPX) values, which are arbitrary units on a log2 scale, where a one-unit increase in NPX corresponds to a two-fold increase in protein concentration. The limit of detection (LOD) for each protein was determined based on three times the standard deviation (SD) above the NPX value of the negative controls in each run. In this study, a total of 183 different proteins were measured, where each panel included 92 proteins involved in diverse biological processes and 4 technical controls. Analyses were performed at the Clinical Biomarker Facility at SciLifeLab, Uppsala University.

### Data analysis

Data analyses were performed using R software (www.r-project.org). Normalized protein expression (NPX) values were used for analysis because they tend to follow a normal distribution. Samples from malaria infected group and the controls were randomized within the plates to minimize variability between plates. The inter-plate variation was further normalized for each protein in each plate by adding a z-score factor, which was calculated as follows: factor = (actual value - median of all samples) / standard deviation. To compare malaria cases and controls, NPX values were adjusted if a significant effect (adjusted p value < 0.05) for age on protein levels was found by linear regression in both the control and malaria groups.

For univariate analysis, the difference in protein levels between the malaria and non-malaria groups was examined using linear regression, using age and gravida as covariates. The dependent variable was NPX, and the independent variables were malaria status. The likelihood ratio test was used to assess the significance of the difference. The statistical significance of the differences between two groups was determined for continuous variables using the Mann–Whitney–Wilcoxon test. For categorical variables, the Chi-square or Fisher’s exact test was used. The analysis of variance (ANOVA) test was used to compare more than two groups. The paired Mann–Whitney–Wilcoxon test was used to compare different malaria groups. Spearman’s rank rho was used to test the correlation coefficients or collinearity between each protein marker and other continuous variables. The Bonferroni–Dunn post hoc test was used to correct for multiple testing by adjusting the p-values for false discovery rate. A difference was considered significant if the q-value (adjusted p-value) was less than 0.05.

Additionally, elastic-net penalized logistic regression (ENLR) was used to identify a combination of analytes that could improve the discrimination between cases and controls. ENLR applies a penalty to the regression coefficients and finds groups of correlated variables. The optimal penalization proportion α was determined using grid search with a 10-fold cross-validation. The optimal tuning parameter λ was determined as the mean value of 100 iterative lambda values that minimized the model’s deviance. The regression coefficients were used to assess the contribution of individual proteins to the case-control discrimination. The ENLR model was estimated using the R package glmnet [19]. The regression coefficients for the selected proteins were then calculated by rerunning ENLR with only proteins with non-zero coefficients after 10 repetitions. To determine the number of proteins required to predict PAM, ROC curves were generated while adding one more protein at a time and then compared with the first ROC curve. This process was repeated until none of the ROC curves showed significant improvement.

## Results

### Protein reactivity

PEA is optimal for quantification of the levels of soluble plasma proteins in small sample volumes. Here, to identify proteins that might indicate *P. falciparum* infection during pregnancy, we probed the Olink Target 96 Inflammation and Olink Target 96 Cardiovascular II Panels (**Table S1**) using serum samples collected from 50 women, who were sampled at three different timepoints during pregnancy (**Table 1**). The initial sampling was performed at 12–18 weeks of gestation and two more samples were collected one and two months before delivery. Twelve (12) women (median age: 24 years, range: 20–29 years) had malaria infection that was detectable using a rapid diagnostic test. The women in the different groups did not differ significantly in terms of age, gravidity, hemoglobin levels, and gestational age.

**Table 1.**
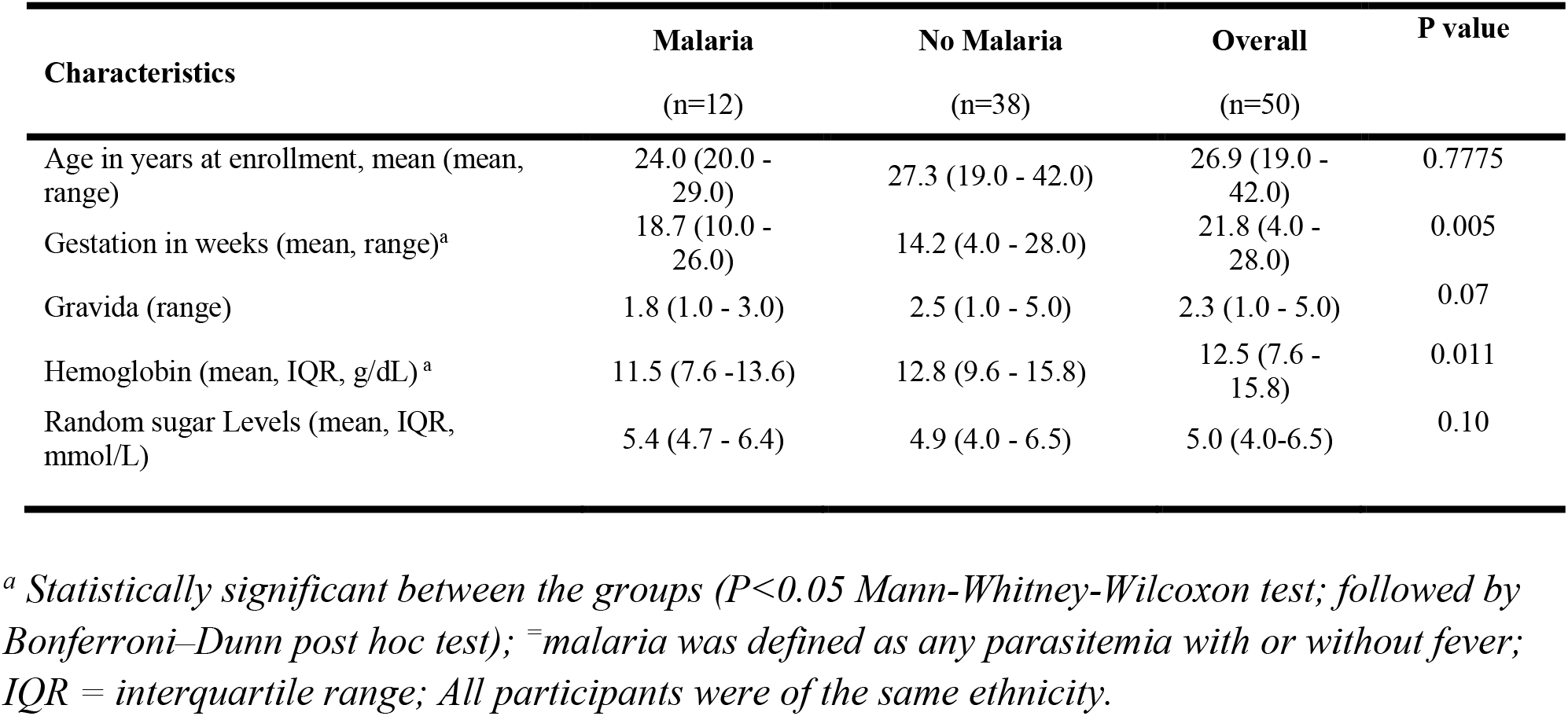
Characteristics of the women assessed in this study.

| Characteristics | Malaria<br>(n=12) | No Malaria<br>(n=38) | Overall<br>(n=50) | P value |
| --- | --- | --- | --- | --- |
| Age in years at enrollment, mean (mean, range) | 24.0 (20.0 - 29.0) | 27.3 (19.0 - 42.0) | 26.9 (19.0 - 42.0) | 0.7775 |
| Gestation in weeks (mean, range)<sup>a</sup> | 18.7 (10.0 - 26.0) | 14.2 (4.0 - 28.0) | 21.8 (4.0 - 28.0) | 0.005 |
| Gravida (range) | 1.8 (1.0 - 3.0) | 2.5 (1.0 - 5.0) | 2.3 (1.0 - 5.0) | 0.07 |
| Hemoglobin (mean, IQR, g/dL)<sup>a</sup> | 11.5 (7.6 - 13.6) | 12.8 (9.6 - 15.8) | 12.5 (7.6 - 15.8) | 0.011 |
| Random sugar Levels (mean, IQR, mmol/L) | 5.4 (4.7 - 6.4) | 4.9 (4.0 - 6.5) | 5.0 (4.0-6.5) | 0.10 |

<sup>a</sup> Statistically significant between the groups ( $P < 0.05$ Mann-Whitney-Wilcoxon test; followed by Bonferroni–Dunn post hoc test); <sup>°</sup>malaria was defined as any parasitemia with or without fever; IQR = interquartile range; All participants were of the same ethnicity.

### Protein reactivity and the characteristic of the sampled women

Protein reactivity to the proteomic panels was measured at three different visits per participant. The pattern of expression profiles varied considerably among the volunteers, with most proteins being present at different time points (first, second and third trimester) for most women. However, in about 10% of the women, less than 20 proteins were detected (**Figure 1**).

**Figure 1:**
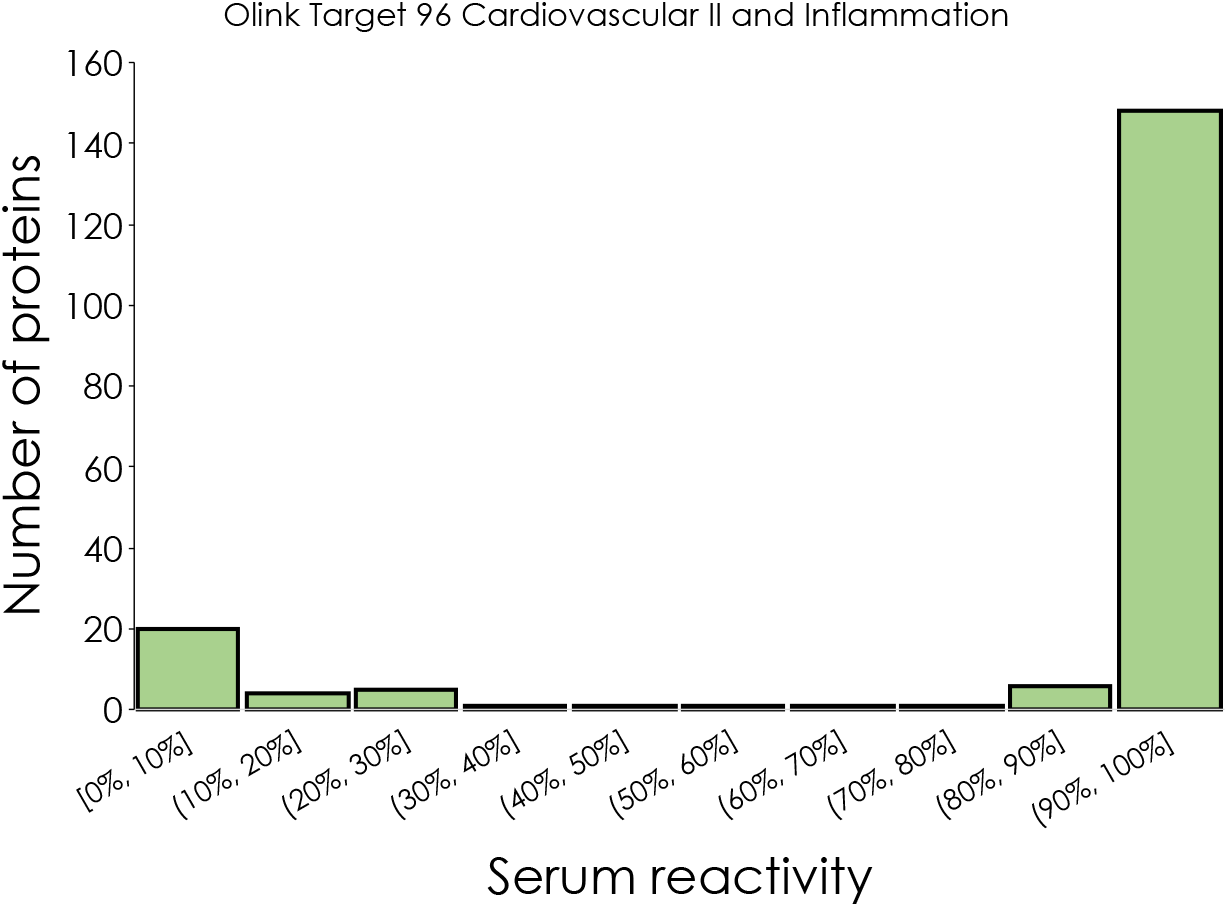
The pattern of plasma proteins expression among pregnant women volunteers assessed from western Kenya.

To assess if there was any relationship between serum protein levels and participant characteristics, such as age, gravida, and gestational age, we conducted a correlation analysis to each of these parameters against the NPX for each protein. Although there was no correlation between protein levels and participants’ age, overall plasma protein levels were higher in samples from primigravida (first-time pregnant) women when compared with multigravida women (who have had 2– 5 pregnancies), although the difference did not reach statistical significance (Table S2).

We further assessed the relationship between markers of morbidity, including hemoglobin and random blood sugars, and expressed plasma proteins. We observed that 19 proteins were significantly negatively correlated with hemoglobin levels, and 9 proteins with random blood sugar levels (unadjusted P < 0.05). However, the correlations did not remain significant after False Discovery Rate (FDR) adjustments. Our findings suggest that these proteins may have a role in malaria infection or that they may be involved in the development and/or progression of pregnancy complications, such as anemia and/or gestational diabetes.

### Differential protein expression in malaria vs no malaria women

Next, we sought to identify differential protein expression in malaria vs non-malaria women. This analysis showed that 48 proteins were significantly differentially expressed in malaria vs no malaria women, at least in one of the three clinic visits (**Table S2**). Notably, IgG Fc receptor IIb (Uniprot ID: P31994) and HO-1 (P09601) were consistently highly expressed in malaria-positive samples when compared with malaria-negative samples, while NRTN (Q99748) and IL-20 (Q9NYY1) were consistently highly expressed in sera from malaria-negative women (**Figure 2**). This data suggest that malaria infection may downregulate or upregulate various proteins.

**Figure 2:**
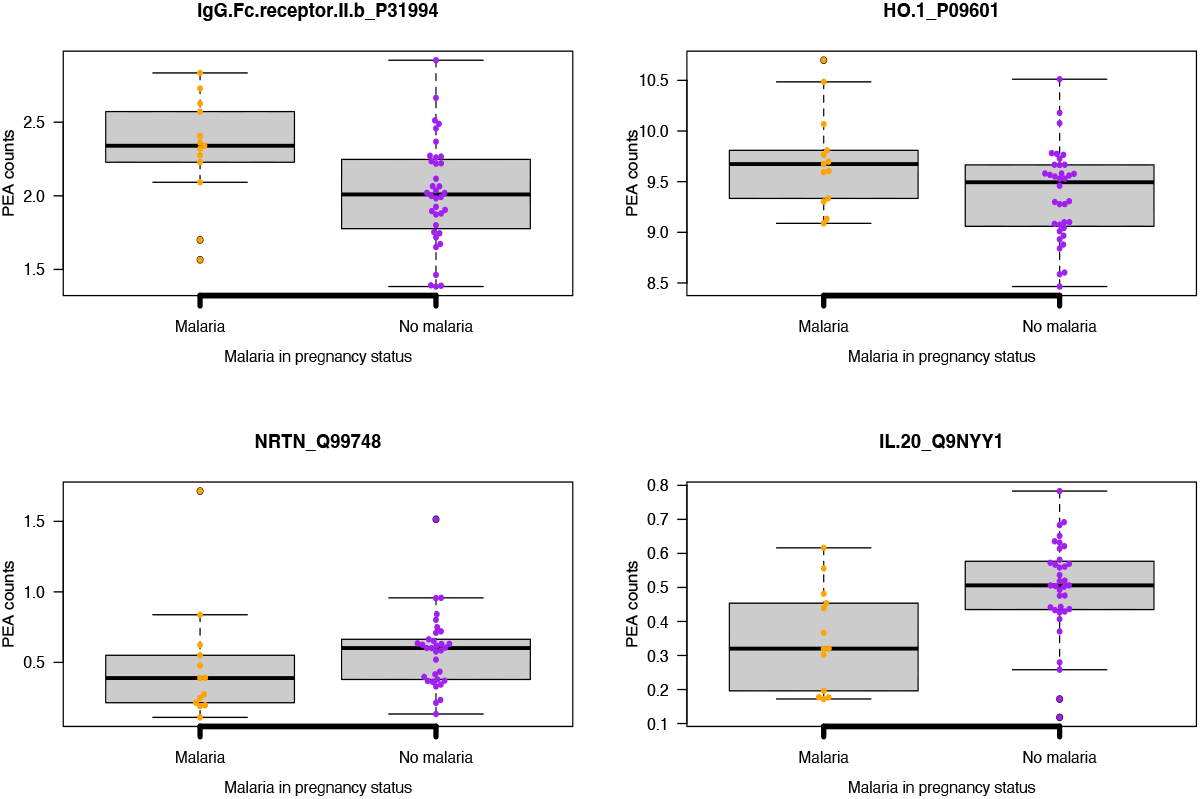
Differential protein expression in malaria-infected vs non malaria-infected women. The title represent protein Uniprot acronym and ID. Relative PEA counts are derived from NPX/ LOD to allow protein-to-protein comparison.

We therefore sought to assess if the levels of these proteins fluctuated during the antepartum period using samples collected at different pregnancy timepoints. Except for IL-20, whose serum levels reduced during the follow-up period, there was no significant change in the other proteins, suggesting that the initial induction by malaria infection did not alter the levels of these proteins during the follow-up period. The top differentially expressed proteins are presented in **Figure 3**, and the full protein list is presented in Supplementary **Tables S3**.

**Figure 3:**
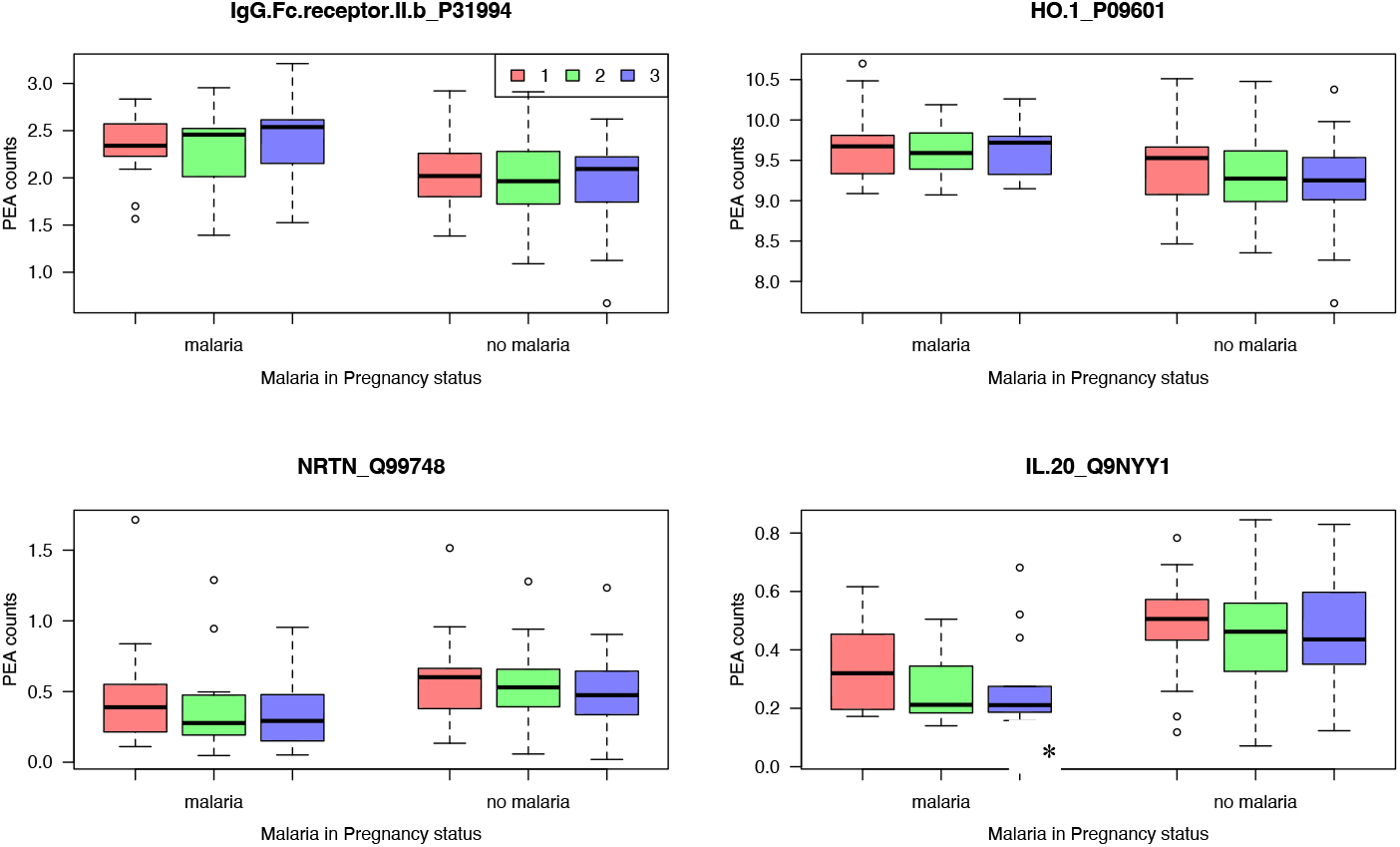
Kinetics of protein expression in malaria-infected vs non-malaria-infected women at three time points during the pregnancy – 1^st^ (red), 2^nd^ (green) and 3^rd^ Visit (blue). * Represent statistical difference between hospital visits, (Kruskal Wallis, P < 0.005).

### Protein levels could distinguish malaria cases vs controls

We also assessed whether a combination of serum proteins could distinguish malaria cases vs controls. This analysis revealed varying predictive performance among the four proteins we examined in relation to malaria status. IL-20 (Q9NYY1) protein exhibited the highest discriminatory power (AUC = 0.815), indicating a strong association with malaria status **(Figure 4)**. This suggests that IL-20 expression levels serve as a promising biomarker for malaria diagnosis or prognosis. IgG Fc receptor IIb (Uniprot ID: P31994) protein demonstrated moderate discriminatory ability (AUC = 0.717), suggesting a notable but less pronounced association with malaria status compared to IL-20. Similarly, HO-1 (P09601) protein also displayed moderate discriminatory power (AUC = 0.729), indicating some association with malaria status, although not as strong as IL-20. NRTN (Q99748) protein, with the lowest AUC value (AUC = 0.635), showed weaker discriminatory ability compared to the other proteins. Although NRTN expression levels may still have some association with malaria status, the association is less pronounced compared to IL-20, IgG Fc receptor IIb, and HO-1.

**Figure 4:**
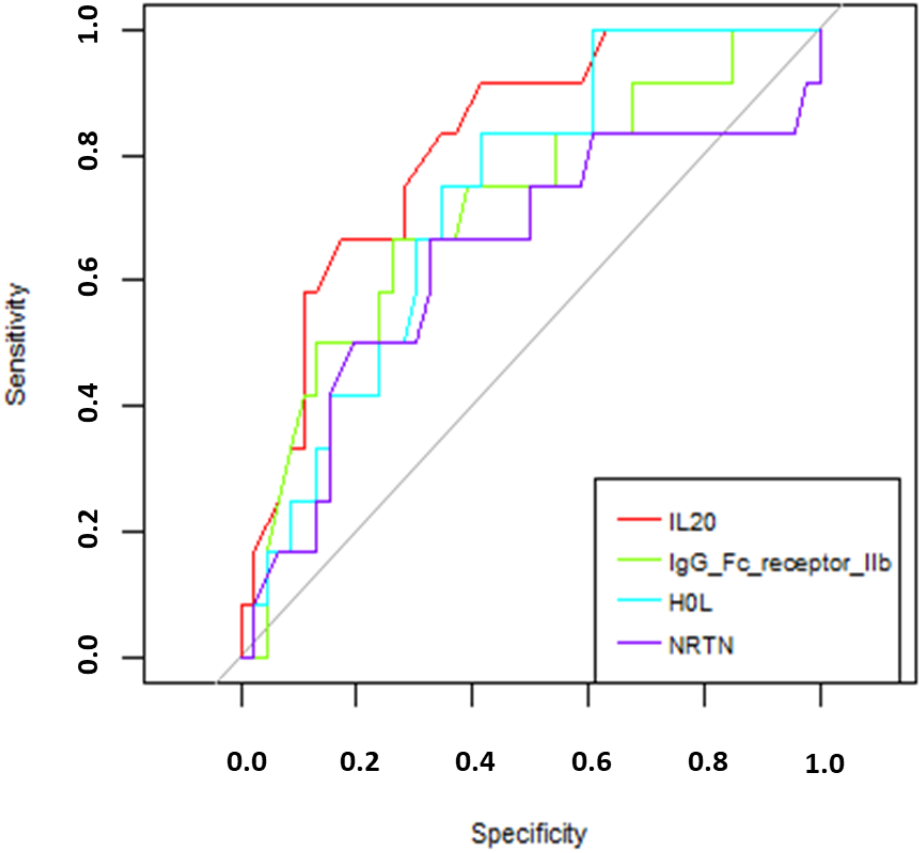
ROC curves for different protein expression levels in malaria-vs non-malaria-infected women. Each protein has been assigned a colour code, as indicated in the legend.

## Discussion

Accurate tools for diagnosing malaria during pregnancy are urgently needed to guide effective clinical interventions [20]. In this study, we aimed to identify potential markers of malaria infection by using high-throughput PEA technology. We observed that IgG Fc receptor IIb (Uniprot ID, P31994), HO-1 (P09601), NRTN (Q99748) and IL-20 (Q9NYY1) were differentially expressed in sera from malaria-infected pregnant women vs from non-malaria-infected controls. Moreover, these proteins may also highlight processes that underlie malaria pathophysiology during pregnancy.

IgG Fc receptor IIb (FCGR2B, Uniprot ID, P31994), a low affinity immunoglobulin gamma Fc region receptor, has multiple isoforms in macrophages, lymphocytes, and IgG-transporting placental epithelium [21]. It is the only inhibitory Fc receptor that controls many aspects of immune and inflammatory responses [22]. Variations in the gene encoding this protein, such as loss-of-function polymorphism, have implications for autoimmunity and infection, including severe malaria. FCGR2B is involved in several processes, including Fc receptor-mediated immune complex endocytosis, nervous system development, and immune responses [23]. Other studies have identified FCGR2B as a marker of human metastatic melanoma, where its expression impairs the tumor susceptibility to FcgammaR-dependent innate effector responses [24]. With the availability of monoclonal antibodies against FCGR2B [25], studies are needed to evaluate its potential as a biomarker of PAM.

We also identified HO-1 (heme oxygenase-1) as abundantly expressed in PAM cases. HO-1 plays a crucial role in heme degradation and cellular responses to stress [26]. During pregnancy, HO-1 maintains a balanced immune response and supports healthy placental development [27]. Evidence from a mouse model [27] indicates that it helps protect against oxidative stress and inflammation, which are often elevated in pregnancy-associated conditions. We hypothesize that in the context of pregnancy-associated malaria, HO-1 may play a role in mitigating the adverse effects of the infection by reducing oxidative stress and inflammation.

Two other proteins that decreased in PAM cases, Neurturin (NRTN) and Interleukin-20 (IL-20), deserve further evaluation. NRTN has been implicated in supporting healthy placental growth and development. It is also involved in the regulation of blood vessel formation and trophoblast invasion, which are critical for successful pregnancy outcomes [28]. IL-20 plays a critical role in the development and maintenance of the placenta, as well as in the regulation of trophoblast invasion, angiogenesis and vascular remodeling [29]. Thus, IL-20 downregulation during PAM might contribute to poor pregnancy outcomes. Indeed, disruptions in IL-20 signaling have been associated with adverse pregnancy outcomes [30]. In the context of pregnancy-associated malaria, IL-20 may modulate the inflammatory response and placental immune tolerance, potentially impacting the severity and outcomes of the infection. These molecules need further evaluation.

The choice of two Olink panels, Inflammation and Cardiovascular II, provided a wide repertoire of proteins with a critical role during pregnancy or with diagnostic capability for assessing malaria infection of the placenta. However, the panels could have excluded other potentially important proteomic targets, and future studies should aim to include a more diverse range of proteins. This study involved a relatively small sample size and was based on a single study center. Hence, these observations warrant further validation using larger, multicenter malaria-in-pregnancy cohorts. Because this study is correlative, future studies will also seek to experimentally validate the involvement of the identified factors in malaria during pregnancy and the underlying mechanisms, which may uncover additional diagnostic and therapeutic strategies.

In conclusion, this study identified four proteins that were differentially regulated between malaria and non-malaria women. Through further investigation, expression of these proteins could potentially be validated for diagnosis of pregnancy-associated malaria.

## Data Availability

All data produced in the present study are available upon reasonable request to the authors.

## Acknowledgements

We appreciate the study volunteers from Webuye County Hospital and thank the research teams from Mount Kenya University for their technical assistance in obtaining and processing the field samples.

## Authors contribution

BNK, MKM, MV and JG conceived and designed experiments. FMK, HW, JM, and RG conducted experiments. BNK, RG, FMK, and HW analyzed the data. BNK, FMK, and JG wrote the manuscript. All authors discussed and edited the manuscript.

## References

[1] Rogerson SJ, Mwapasa V, Meshnick SR. Malaria in pregnancy: linking immunity and pathogenesis to prevention. Am J Trop Med Hyg 2007;77:14–22.

[2] Brabin BJ. An analysis of malaria in pregnancy in Africa. Bull World Health Organ 1983;61:1005–16.

[3] Brabin BJ, Johnson PM. Placental malaria and pre-eclampsia through the looking glass backwards? J Reprod Immunol 2005;65:1–15. 10.1016/j.jri.2004.09.006.

[4] Roberts JM, Lain KY. Recent Insights into the Pathogenesis of Pre-eclampsia. Placenta 2002;23:359–72. 10.1053/plac.2002.0819.

[5] Jena MK, Sharma NR, Petitt M, Maulik D, Nayak NR. Pathogenesis of Preeclampsia and Therapeutic Approaches Targeting the Placenta. Biomolecules 2020;10. 10.3390/biom10060953.

[6] Gueneuc A, Deloron P, Bertin GI. Usefulness of a biomarker to identify placental dysfunction in the context of malaria. Malar J 2017;16:1–7. 10.1186/s12936-016-1664-0.

[7] Acharya P, Garg M, Kumar P, Munjal A, Raja KD. Host-parasite interactions in human malaria: Clinical implications of basic research. Front Microbiol 2017;8:1–16. 10.3389/fmicb.2017.00889.

[8] Unger HW, Ome-Kaius M, Wangnapi RA, Umbers AJ, Hanieh S, Suen CSNLW, et al. Sulphadoxine-pyrimethamine plus azithromycin for the prevention of low birthweight in Papua New Guinea: A randomised controlled trial. BMC Med 2015;13:1–16. 10.1186/s12916-014-0258-3.

[9] Wik L, Nordberg N, Broberg J, Björkesten J, Assarsson E, Henriksson S, et al. Proximity Extension Assay in Combination with Next-Generation Sequencing for High-throughput Proteome-wide Analysis. Mol Cell Proteomics 2021;20:100168. 10.1016/j.mcpro.2021.100168.

[10] Dayon L, Cominetti O, Affolter M. Proteomics of human biological fluids for biomarker discoveries: technical advances and recent applications. Expert Rev Proteomics 2022;19:131–51. 10.1080/14789450.2022.2070477.

[11] Assarsson E, Lundberg M, Holmquist G, Björkesten J, Thorsen SB, Ekman D, et al. Homogenous 96-plex PEA immunoassay exhibiting high sensitivity, specificity, and excellent scalability. PLoS One 2014;9. 10.1371/journal.pone.0095192.

[12] Allan-Blitz L-T, Akbari O, Kojima N, Saavedra E, Chellamuthu P, Denny N, et al. Unique immune and inflammatory cytokine profiles may define long COVID syndrome. Clin Exp Med 2023;23:2925–30. 10.1007/s10238-023-01065-6.

[13] Angerfors A, Brännmark C, Lagging C, Tai K, Månsby Svedberg R, Andersson B, et al. Proteomic profiling identifies novel inflammation-related plasma proteins associated with ischemic stroke outcome. J Neuroinflammation 2023;20:1–11. 10.1186/s12974-023-02912-9.

[14] Davies MPA, Sato T, Ashoor H, Hou L, Liloglou T, Yang R, et al. Plasma protein biomarkers for early prediction of lung cancer. EBioMedicine 2023;93:1–11. 10.1016/j.ebiom.2023.104686.

[15] Guterstam YC, Acharya G, Schott K, Björkström NK, Gidlöf S, Ivarsson MA. Immune cell profiling of vaginal blood from patients with early pregnancy bleeding. Am J Reprod Immunol 2023;90:1–13. 10.1111/aji.13738.

[16] Platt A, Obala AA, MacIntyre C, Otsyula B, Meara WPO. Dynamic malaria hotspots in an open cohort in western Kenya. Sci Rep 2018;8:1–11. 10.1038/s41598-017-13801-6.

[17] Peters PJ, Thigpen MC, Parise ME, Newman RD. Safety and toxicity of sulfadoxine/pyrimethamine: implications for malaria prevention in pregnancy using intermittent preventive treatment. Drug Saf 2007;30:481–501. 10.2165/00002018-200730060-00003.

[18] Dixon JRJ. The International Conference on Harmonization Good Clinical Practice guideline. Qual Assur 1998;6:65–74. 10.1080/105294199277860.

[19] Fredriksson S, Gullberg M, Jarvius J, Olsson C, Pietras K, Gústafsdóttir SM, et al. Protein detection using proximity-dependent DNA ligation assays. Nat Biotechnol 2002;20:473–7. 10.1038/nbt0502-473.

[20] Friedman JH, Hastie T, Tibshirani R. Regularization Paths for Generalized Linear Models via Coordinate Descent. J Stat Softw 2010;33:1–22. 10.18637/jss.v033.i01.

[21] Ding XC, Incardona S, Serra-Casas E, Charnaud SC, Slater HC, Domingo GJ, et al. Malaria in pregnancy (MiP) studies assessing the clinical performance of highly sensitive rapid diagnostic tests (HS-RDT) for Plasmodium falciparum detection. Malar J 2023;22:1–15. 10.1186/s12936-023-04445-1.

[22] Stuart SG, Simister NE, Clarkson SB, Kacinski BM, Shapiro M, Mellman I. Human IgG Fc receptor (hFcRII; CD32) exists as multiple isoforms in macrophages, lymphocytes and IgG-transporting placental epithelium. EMBO J 1989;8:3657–66. 10.1002/j.1460-2075.1989.tb08540.x.

[23] Smith KGC, Clatworthy MR. Europe PMC Funders Group Fc γ RIIB in autoimmunity and infection: evolutionary and therapeutic implications 2014;10:328–43. 10.1038/nri2762.Fc.

[24] Bournazos S, Woof JM, Hart SP, Dransfield I. Functional and clinical consequences of Fc receptor polymorphic and copy number variants. Clin Exp Immunol 2009;157:244–54. 10.1111/j.1365-2249.2009.03980.x.

[25] Cassard L, Cohen-Solal JFG, Fournier EM, Camilleri-Broët S, Spatz A, Chouaïb S, et al. Selective expression of inhibitory Fcγ receptor by metastatic melanoma impairs tumor susceptibility to IgG-dependent cellular response. Int J Cancer 2008;123:2832–9. 10.1002/ijc.23870.

[26] Veri MC, Gorlatov S, Li H, Burke S, Johnson S, Stavenhagen J, et al. Monoclonal antibodies capable of discriminating the human inhibitory Fcγ-receptor IIB (CD32B) from the activating Fcγ-receptor IIA (CD32A): Biochemical, biological and functional characterization. Immunology 2007;121:392–404. 10.1111/j.1365-2567.2007.02588.x.

[27] Ozen M, Zhao H, Kalish F, Yang Y, Jantzie LL, Wong RJ, et al. Inflammation-induced alterations in maternal-fetal Heme Oxygenase (HO) are associated with sustained innate immune cell dysregulation in mouse offspring. PLoS One 2021;16:1–18. 10.1371/journal.pone.0252642.

[28] Schumacher A, Zenclussen AC. Effects of heme oxygenase-1 on innate and adaptive immune responses promoting pregnancy success and allograft tolerance. Front Pharmacol 2015;5:1–9. 10.3389/fphar.2014.00288.

[29] Morel L, Domingues O, Zimmer J, Michel T. Revisiting the Role of Neurotrophic Factors in Inflammation. Cells 2020;9. 10.3390/cells9040865.

[30] Menon R, Ismail L, Ismail D, Merialdi M, Lombardi SJ, Fortunato SJ. Human fetal membrane expression of IL-19 and IL-20 and its differential effect on inflammatory cytokine production. J Matern Neonatal Med 2006;19:209–14. 10.1080/14767050500440986.

